# Who Does What to Whom? Graph Representations of Action-Predication in Speech Relate to Psychopathological Dimensions of Psychosis

**DOI:** 10.1101/2022.03.18.22272636

**Authors:** Amir H. Nikzad, Yan Cong, Sarah Berretta, Katrin Hänsel, Sunghye Cho, Sameer Pradhan, Leily Behbehani, Danielle DeSouza, Mark Y. Liberman, Sunny X. Tang

**Affiliations:** Feinstein Institutes for Medical Research, Zucker Hillside Hospital, Department of Psychiatry; Yale University, Department of Laboratory Medicine; University of Pennsylvania, Linguistic Data Consortium; Winterlight Labs, Inc.; Stanford University, Department of Neurology & Neurological Sciences

## Abstract

Graphical representations of speech generate powerful computational measures related to psychosis. Previous studies have mostly relied on structural relations between words as the basis of graph formation, i.e. connecting each word to the next in a sequence of words. Here, we introduced a method of graph formation grounded in semantic relationships by identifying elements that act upon each other (action relation) and contents of those actions (predication relation). Speech from picture description and open-ended narrative tasks were collected from a cross-diagnostic group of healthy volunteers and people with psychotic as well as non-psychotic disorders. Recordings were transcribed and underwent automated language processing, including semantic role labeling to identify action and predication relations. Structural and semantic graph features were computed using static and dynamic (moving-window) techniques. Compared to structural graphs, semantic graphs were more highly correlated with dimensional psychosis symptoms. Dynamic features also outperformed static features, and samples from picture descriptions were more informative than narrative responses for psychosis diagnoses and symptom dimensions. Overall, semantic graphs capture unique and clinically meaningful information about psychosis and related symptom dimensions. These features, particularly when derived from semi-structured tasks using dynamic measurement, are meaningful additions to the repertoire of computational linguistic methods in psychiatry.

## INTRODUCTION

Disturbances in speech have been recognized as a key component of both positive and negative symptoms in psychosis [1].^1^ Increasingly, speech phenotypes in psychosis can be objectively and reliably measured through automated language analysis for detection and prediction of psychotic disorders [2-7]. Speech graphs, derived from transcribed text, have shown the ability to accurately quantify language disorganization and impoverishment. There are significant relationships between graph features and key psychosis phenotypes, including thought disorder, cognition, global functioning, and brain connectivity changes [8]. Speech graphs are network representations of discourse which treat linguistic elements (words, lexemes, etc.) as nodes and relationships among those elements as the bridging links (edges) [9]. Generally, relationships among linguistic elements may be structural (based on the relative locations of the words, e.g., occurring in sequence or co-occurrence in the same utterance) or semantic (based on the meaning of the utterance, e.g., entity A is acting upon entity B). Quantitative measures of the size, connectedness, and organizational structure of the speech graphs can then be calculated [10]. For example, the size of the graph can be quantified by number of nodes and edges as well as measures of internal distances such as network diameter and average shortest path length. The connectedness of the graph is reflected in average degrees, graph density, and size of largest connected component. The degree of organization in the graph can be measured by statistical similarity of graph features to randomly generated graphs of the same size.

Sequential speech graphs of individuals with psychosis spectrum disorders (PS+) have been characterized as being smaller, less connected and more disorganized than individuals without psychosis spectrum disorders (PS-). In their pioneering study, Mota and colleagues showed that the PS+ speech graphs exhibited fewer nodes and edges, lower average degrees and smaller connected components compared to those of healthy controls and patients with mania [11]. PS+ speech graphs were also found to have lower average shortest path length and network diameter per fixed word lengths, reflecting shorter internal distances [12]. A subsequent study comparing the size of connected components in speech graphs with that of randomly generated graphs of the same size revealed that PS+ graphs have more random-like organization compared to PS- [13]. Palaniyappan et al. (2019) further showed that the graph measures of connectedness (size of connected component) and organization (size of connected component divided by that of random graphs) are associated with disorganized and impoverished thought disorders as well as clinical measures of schizophrenic symptoms and biological measures of neural connectivity.

However, sequential relationships among words are vulnerable to non-psychopathologic factors such as stylistic preferences of the speaker, passive or active voice, and different grammatical structures in different languages.On the other hand, semantic relationships are tied to the underlying concepts being expressed, and are consistent across languages and speaking style. For instance, in “The dog chased the cat” and “The cat was chased by the dog,” the same semantic content is expressed with distinct word sequences. Furthermore, there is evidence that thought disorder is related to disruptions in semantic networks [14]. Thus, graphs which utilize the core semantic content may be a more direct method for representing disrupted brain circuits in psychosis. Here, we attempt to build speech graphs upon two semantic relations which have been proposed as universal linguistic relationships [15]: 1) Action, which links the actor of the underlying event to its undergoers (dog → cat), and 2) predications, which link the predicate of the utterance to its arguments (chase → dog and cat). These relationships capture the core semantic meaning of the utterance: who does what to whom? Figure 1-A illustrates the structures of sequential and action-predication graphs.

**Figure 1.**
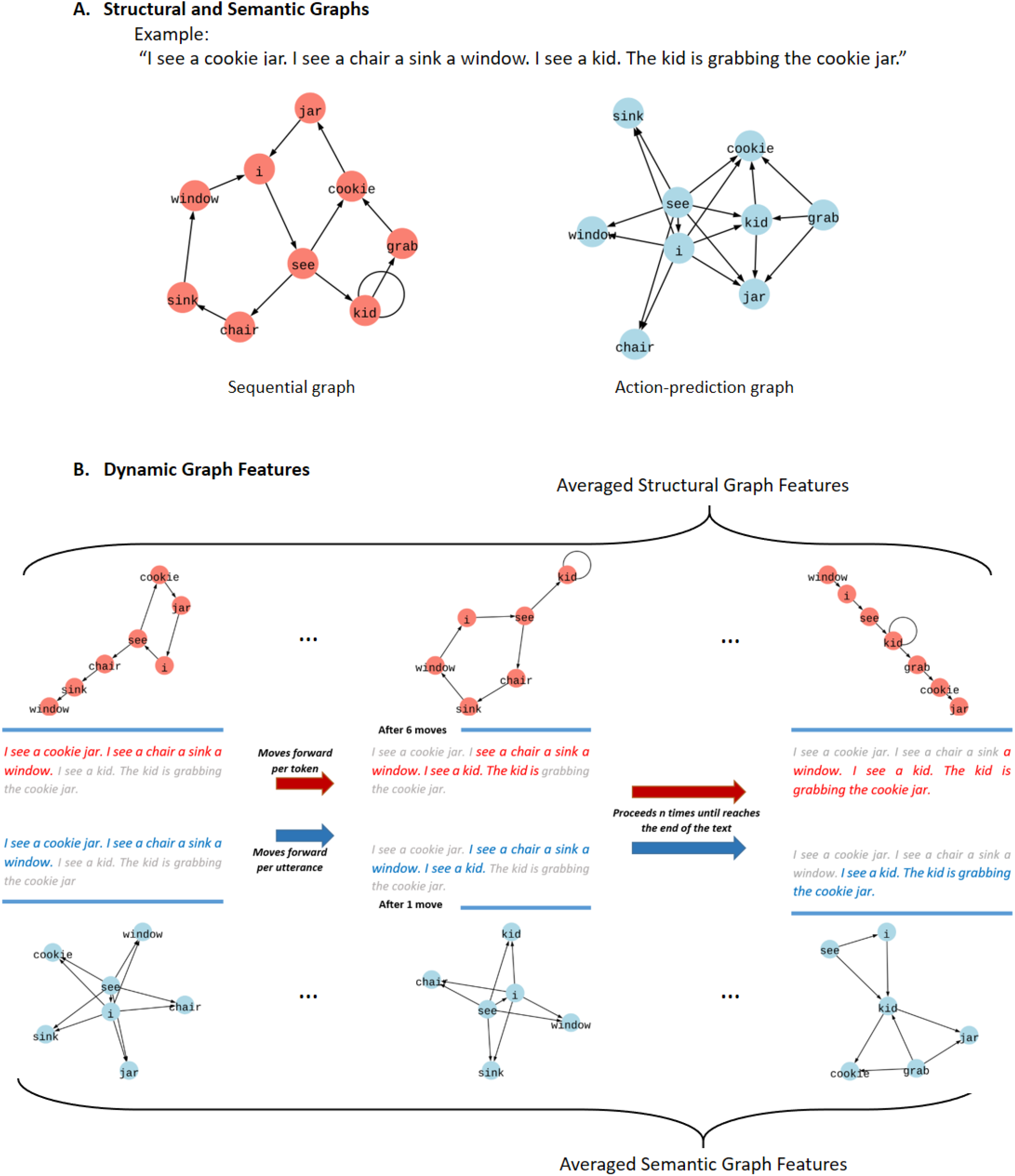
Speech Graph Methodology. (A) Structural and semantic graph representations of a given text are illustrated. The structural representation is produced based on sequential relations between lemmatized content words (e.g. I→see→cookie→jar, etc.). The semantic representation is produced by connecting elements that act upon each other (e.g. I→chair; kid→cookie jar), and linking verb-predicates to their arguments (e.g. see→I; see→chair; grab→kid; grab→cookie jar). (B) Dynamic graph features are computed by sliding a window of fixed-length throughout each sample to producing *n* instances of graph representations. Subsequently, each graph feature is calculated as the mean value of *n* features each belonging to a particular instance. Successive sequential graphs progress one word at a time in windows of 30 words, and semantic graphs slide one utterance at a time in windows of 3 utterances.

In this paper, we aim at 1) introducing a new way to produce semantic speech graphs based on the two universal linguistic relations of action and predication, 2) verifying validity of size, connectedness and organization as three non-redundant domains of speech graph features, 3) comparing semantic and structural graph measures of size, connectedness and organization in their associations with the presence of psychosis, and 4) examining the relationship between graph features and clinically-rated dimensions of psychosis symptoms. The overall goal is to operationalize semantic speech graph methodology with respect to studying language disturbance in psychosis and guide subsequent studies.

## RESULTS

### Participant Characteristics

In total, speech samples of 205 and 201 participants were collected and transcribed for picture description and open-ended narrative tasks respectively, corresponding to 81 PS+ and 124 PS-participants (Table 1). On average, picture descriptions included 110 ± 66 words and narrative responses included 162 ± 121 words; word counts were not significantly different between PS+ and PS- (p-values were 0.310 and 0.051 for open-ended narrative and picture description tasks respectively). As expected, participants with psychosis scored significantly higher in overall psychosis symptoms, negative symptoms, and demonstrated significantly more abnormal speech per clinical ratings.

**Table 1.**
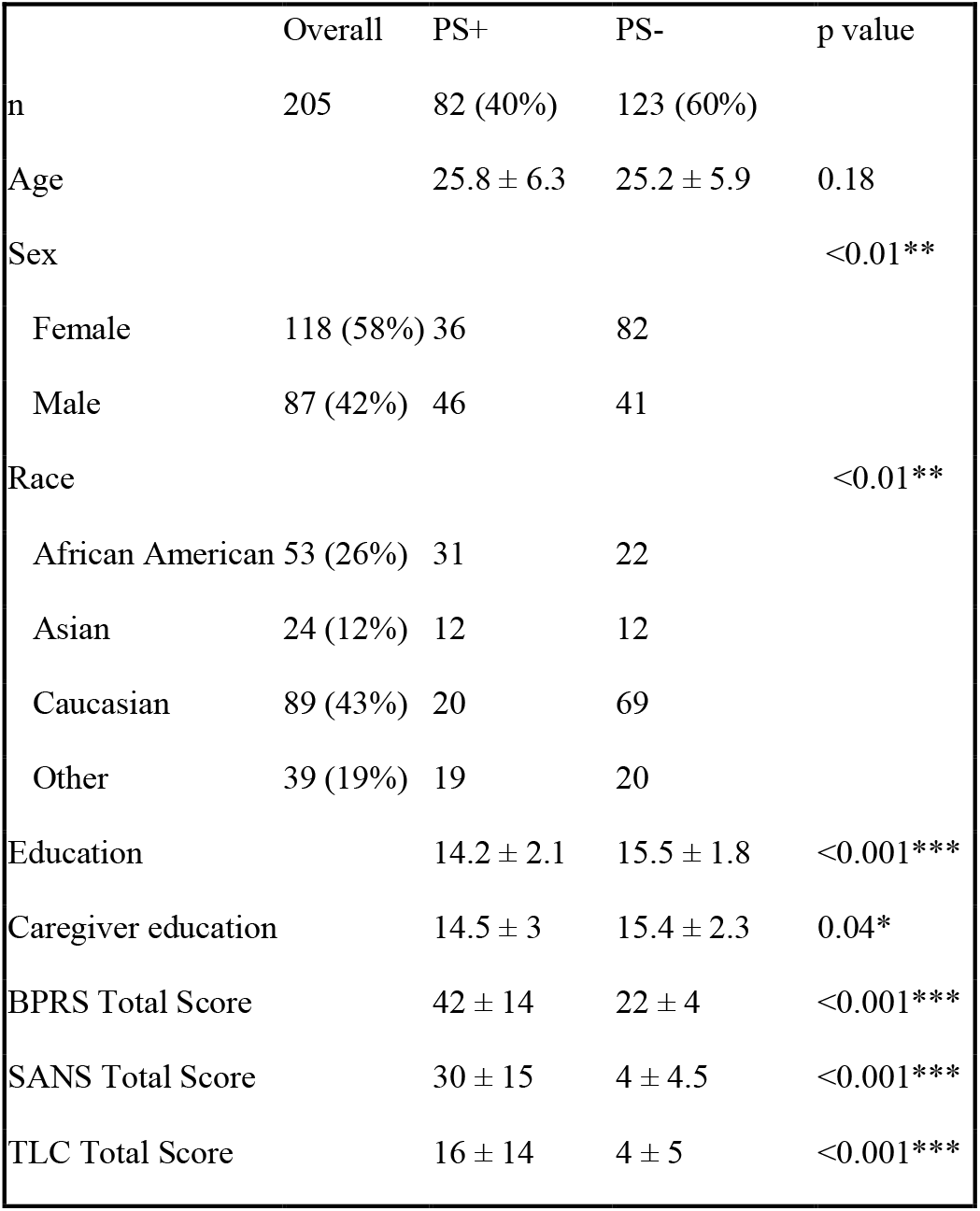
Demographic characteristics of the participants. The significant associations are tagged by asterisks (*=<0.05, **=<0.01, and ***<0.001). PS+: Individuals with psychosis spectrum disorders; PS-: Individuals without psychosis spectrum disorders); BPRS: Brief Psychiatric Rating Scale; SANS: Scale for Assessment of Negative Symptoms; TLC: Scale for the Assessment of Thought, Language and Communication. P-values were calculated using Pearson’s chi-squared test (for sex and race), and Mann-Whitney U test (for age, education, caregiver education, and BPRS, SANS and TLC total scores).

|  | Overall | PS+ | PS- | p value |
| --- | --- | --- | --- | --- |
| n | 205 | 82 (40%) | 123 (60%) |  |
| Age |  | 25.8 ± 6.3 | 25.2 ± 5.9 | 0.18 |
| Sex |  |  |  | <0.01** |
| Female | 118 (58%) | 36 | 82 |  |
| Male | 87 (42%) | 46 | 41 |  |
| Race |  |  |  | <0.01** |
| African American | 53 (26%) | 31 | 22 |  |
| Asian | 24 (12%) | 12 | 12 |  |
| Caucasian | 89 (43%) | 20 | 69 |  |
| Other | 39 (19%) | 19 | 20 |  |
| Education |  | 14.2 ± 2.1 | 15.5 ± 1.8 | <0.001*** |
| Caregiver education |  | 14.5 ± 3 | 15.4 ± 2.3 | 0.04* |
| BPRS Total Score |  | 42 ± 14 | 22 ± 4 | <0.001*** |
| SANS Total Score |  | 30 ± 15 | 4 ± 4.5 | <0.001*** |
| TLC Total Score |  | 16 ± 14 | 4 ± 5 | <0.001*** |

### Speech Graphs Formation and Measurements

The structural graph representations were formed by sequential connection of elements of structural graph entry irrespective of utterance boundaries. The semantic graph representations were created by first tagging verbs, actor-arguments and undergoer-arguments using semantic role labeling, and then combining action relations (actor → undergoer) and predication relations (verb-predicate → actor and undergoer arguments) within each utterance (Figure 1-A). Iterations of the same relationships were captured as the edge weight; the first occurrence was weighted 1 and repetitions added 1 to the edge weight incrementally. We shared our code for the formation of semantic graphs in a public repository (https://github.com/STANG-lab/Semantic-Graphs).

*Utterance Example:*

> The kid is grabbing the cookie jar.
>
> *Structural Graph Connections:*
>
> kid → grab + grab → cookie + cookie → jar
>
> *Semantic Graph Connections:*
>
> grab → kid + grab → cookie jar + kid → cookie jar

Size, connectedness and organization of sequential and action-predication networks were measured by computing relevant graph features. Graph size was quantified with straightforward counts (number of nodes (NN), number of edges (NE)) and internal-distance measures (diameter, average shortest path length between any two nodes (ASPL)). Connectedness was quantified using average weighted degree (i.e., the number of weighted edges for each node; AWD), graph density (realized edges divided by possible edges) and number of nodes in the largest strongly connected component (LSCC). Level of organization in graphs was estimated by computing the z-score of ASPL and LSCC relative to 1000 randomly-generated graphs of the same NN and NE. All calculations were performed in Python using the igraph library [16]. Random graphs were generated using the built-in Erdős– Rényi algorithm [17].

Static graph features were calculated based on the networks of the whole response for each task, and averaged across each task category for each participant (picture description vs. narrative).We complemented usual static graph features with moving-window measures introduced by Mota and colleagues [12]. The length and moving-step of the window were set to 30-tokens length and 1-token step for the sequential graphs based on the best performing models in previous studies which explored this technique [8, 12, 13]. Action-predication networks were calculated for 3-utterance segments and 1-utterance steps because these lengths were the closest equivalents to that of structural graphs, considering the mean sentence length of approximately 10 words. Dynamic graph features were then calculated as the average of graph features over all windows. In total, 36 graph features (n=18 for structural graphs, n=18 for semantic graphs) were analyzed. The full list of graph features is included in Supplemental Table 2.

### The Redundancy of Graph Features

Variance inflation factor (VIF) was computed for different sets of graph features as a measure of their mutual redundancy [18]. Table 2 presents the results of successive VIF comparisons with step-wise increase in the levels of integration of graph features, starting from intra-domain comparison (size vs. connectedness vs. organization) and proceeding to comparisons per mode (static vs. dynamic), type (structural vs. semantic) and task (picture description vs. open-ended). Intra-domain VIF analyses revealed that the information in the internal-distance measures was not redundant with respect to general measures of size. All connectedness measures in both graph types survived in the intra-domain analysis. Dynamic organization measures of sequential graphs were not mutually redundant, but only one of the dynamic organization measures survived in the VIF comparison of the action-predication networks. Increasing the level of integration did not lead to exclusion of one entire domain. Notably, dynamic semantic graph features of all three domains of size, connectedness and organization remained in the final sets in both tasks; this was not observed in other types of graph features.

**Table 2.**
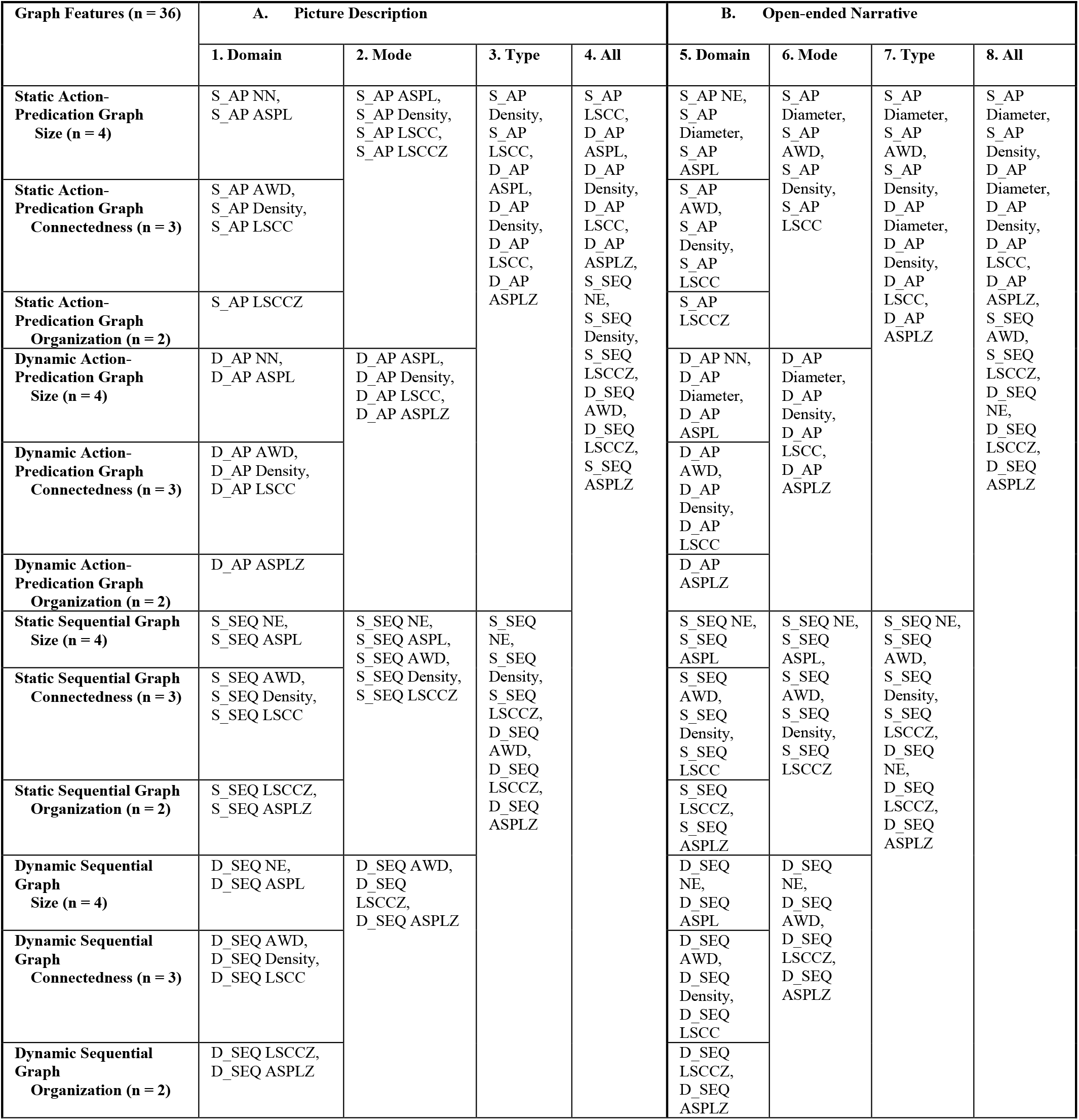
Survived Graph Features in Sequential VIF Comparisons. (A) and (B) correspond to picture description and open-ended narrative tasks, respectively. Columns within each segment accommodate survived graph features. VIF comparison was conducted and features of highest VIF were excluded successively until a set of features all showing VIF < 5 was attained. Survived features were then passed to the next column on right for another comparison on a more integrated level. 1. Domain column shows results of intra-domain comparison. 2. Mode column shows the results for static and semantic features within each graph type. 3. Type column presents the features integrated on graph-type level, i.e. semantic vs structural graph features. 4. All column combines all graph features per each task. For dynamic semantic graphs of both tasks, features belonging to three domains of size, connectedness and organization remained in the final set. Note: S_AP = static action-predication graph feature; D_AP = dynamic action-predication graph feature; S_SEQ = static sequential graph feature; D_SEQ = dynamic sequential graph feature; NN = number of nodes; NE = number of edges; Diameter = graph diameter; ASPL = average shortest path length; AWD = average weighted degree; Density = graph density; LSCC = size of largest strongly connected component; LSCCZ = z-score of LSCC compared to 1000 random graphs; ASPLZ = z-score of ASPL compared to 1000 random graphs. More details on graph features are available in Supplemental Table 2.

### Speech Graph Features and Psychosis

Associations between graph features and presence of psychosis are presented in Table 3. In general, semantic graphs showed a more task-specific behavior compared to structural graphs, with more significant associations in picture descriptions. Structural graphs performed similarly in picture description compared to open-ended narrative tasks. The moving window method enhanced the significance and effect size of the relationships for both graph types and for all three domains of size, connectedness and organization.

**Table 3.** Relationships between Structural and Semantic Graph Features and Psychosis. Graph features are categorized into three domains of size, connectedness and psychosis. For each features the values for picture description task (Picture) and open-ended narrative (Open-Ended) are reported. The significant correlations are tagged by asterisks (*=<0.05, **=<0.01, and ***<0.001). The effect sizes are measured using rank biserial correlation coefficients (RBC). Semantic graphs showed a preference for picture description tasks with higher effect sizes and more significant correlations. Structural graphs showed a task-independent relation with psychosis. In overall, dynamic graph features outperformed static counterparts in both graph types.

| Task |  | Static Action-Predication Graph (S_AP) |  | Dynamic Action-Predication Graph (D_AP) |  | Static Sequential Graph (S_Seq) |  | Dynamic Sequential Graph (D_Seq) |  |
| --- | --- | --- | --- | --- | --- | --- | --- | --- | --- |
|  |  | p value | RBC | p value | RBC | p value | RBC | p value | RBC |
| **A. Size** |  |  |  |  |  |  |  |  |  |
| Number of Nodes (NN) | Picture | <0.05* | 0.19 | <0.01** | 0.26 | <0.01** | 0.22 | <0.001*** | 0.36 |
|  | Open-Ended | 0.09 | 0.14 | <0.01** | 0.23 | 0.06 | 0.15 | <0.001*** | 0.32 |
| Number of Edges (NE) | Picture | <0.05* | 0.20 | <0.001*** | 0.29 | <0.05* | 0.18 | <0.01** | 0.27 |
|  | Open-Ended | 0.12 | 0.13 | <0.05* | 0.20 | 0.14 | 0.12 | <0.001*** | 0.39 |
| Diameter | Picture | <0.05* | 0.17 | <0.05* | 0.21 | <0.05* | 0.26 | <0.001*** | 0.32 |
|  | Open-Ended | 0.86 | 0.01 | 0.65 | 0.04 | <0.01** | 0.27 | <0.01** | 0.24 |
| Average Shortest Path Length (ASPL) | Picture | 0.14 | 0.12 | <0.05* | 0.16 | <0.01** | 0.25 | <0.001*** | 0.30 |
|  | Open-Ended | 0.28 | 0.09 | <0.05* | 0.18 | <0.01** | 0.26 | <0.01** | 0.24 |
| **B. Connectedness** |  |  |  |  |  |  |  |  |  |
| Average Weighted Degree (AWD) | Picture | 0.09 | 0.14 | <0.05* | 0.19 | 0.49 | -0.06 | <0.0001*** | -0.34 |
|  | Open-Ended | 0.9 | -0.01 | 0.35 | 0.08 | 0.17 | -0.11 | <0.01** | -0.27 |
| Density | Picture | 0.31 | -0.08 | 0.08 | -0.15 | <0.001*** | -0.28 | <0.001*** | -0.35 |
|  | Open-Ended | <0.05* | -0.20 | <0.01** | -0.24 | <0.01** | -0.22 | <0.001*** | -0.30 |
| Size of Largest Strongly Connected Component (LSCC) | Picture | 0.08 | 0.14 | <0.01** | 0.22 | <0.01** | 0.21 | 0.06 | 0.16 |
|  | Open-Ended | 0.26 | 0.09 | 0.10 | 0.14 | 0.06 | 0.15 | <0.001*** | 0.30 |
| **C. Organization** |  |  |  |  |  |  |  |  |  |
| LSCC z-score (LSCCZ) | Picture | <0.05* | -0.21 | <0.001*** | -0.29 | <0.001*** | 0.29 | <0.01** | 0.26 |
|  | Open-Ended | 0.19 | -0.11 | 0.09 | -0.14 | <0.01** | 0.26 | <0.001*** | 0.31 |
| ASPL z-score (ASPLZ) | Picture | <0.05* | -0.21 | <0.001*** | -0.28 | 0.92 | -0.01 | <0.01** | 0.27 |
|  | Open-Ended | 0.13 | -0.13 | 0.09 | -0.15 | 0.99 | -0.00 | <0.05* | 0.22 |

#### Size

Speech from PS+ yielded significantly smaller structural and semantic graphs as reflected in multiple correlations between measures of network size and psychosis (Table 3-A). The moving-window technique improved the correlations in both graph types with 0.91 and 0.87 increase in mean RBC for sequential and action-predication graphs respectively. General measures of the network size (NE and NN) manifested the strongest and most significant correlations in psychosis for both graph types and in both tasks.

#### Connectedness

Psychosis was associated with smaller connected components and higher graph densities (Table 3-B). This pattern was observed in both structural and semantic graphs. However, average weighted degrees were higher in the structural graphs of psychotic speech and lower in semantic networks compared to non-psychotic counterparts. The most informative semantic graph features in this domain were dynamic graph density and LSCC for open-ended and picture description tasks respectively. Dynamic density in sequential graphs showed the highest effect size in both tasks (picture description: p<0.001, RBC=-0.35; narrative: p<0.001, RBC=-0.30).

#### Organization

Structural and semantic speech graphs showed different patterns of organization with respect to random graphs with negative mean z-scores in semantic graphs and positive z-scores for structural graphs. This pattern was consistent for both measures (LCCZ and ASPLZ) and in both tasks. These findings suggest that action-predication graphs of speech are organized in smaller connected components with shorter inter-nodal pathways compared to random graphs of the same size. Conversely, sequential graphs produce larger connected components with nodes that were further apart. However, in both cases, PS+ graphs incline toward the more random-like patterns of organizations compared to those of PS-, i.e. exhibiting higher z-scores in semantic graphs and lower z-scores in structural graphs (Table 3-C).

### Speech Graphs and Dimensional Clinical Characteristics

Figure 2-A represents task-wise correlation plots for structural and sematic graph features vs. clinical measures of language disturbance (TLC speech disorganization and speech poverty factors), disease severity (BPRS total score) and psychopathological dimensions (BPRS anxiety/depression, hostility/suspiciousness, though disturbance and withdrawal/psychomotor retardation factors; SANS affective flattening, alogia, avolition, and asociality/anhedonia global scores). In general, graph features generated from the picture description tasks were more closely related to clinical measures than those from narrative tasks. Within the picture description task, the dynamic action-predication graph features outperformed the static features as well as both static and dynamic sequential graph features (Figure 2-A). Among graph measures of the narrative task, a dynamic measure of structural graph organization (D_SEQ LSCCZ) showed the strongest connections with clinical characteristics.

**Figure 2.**
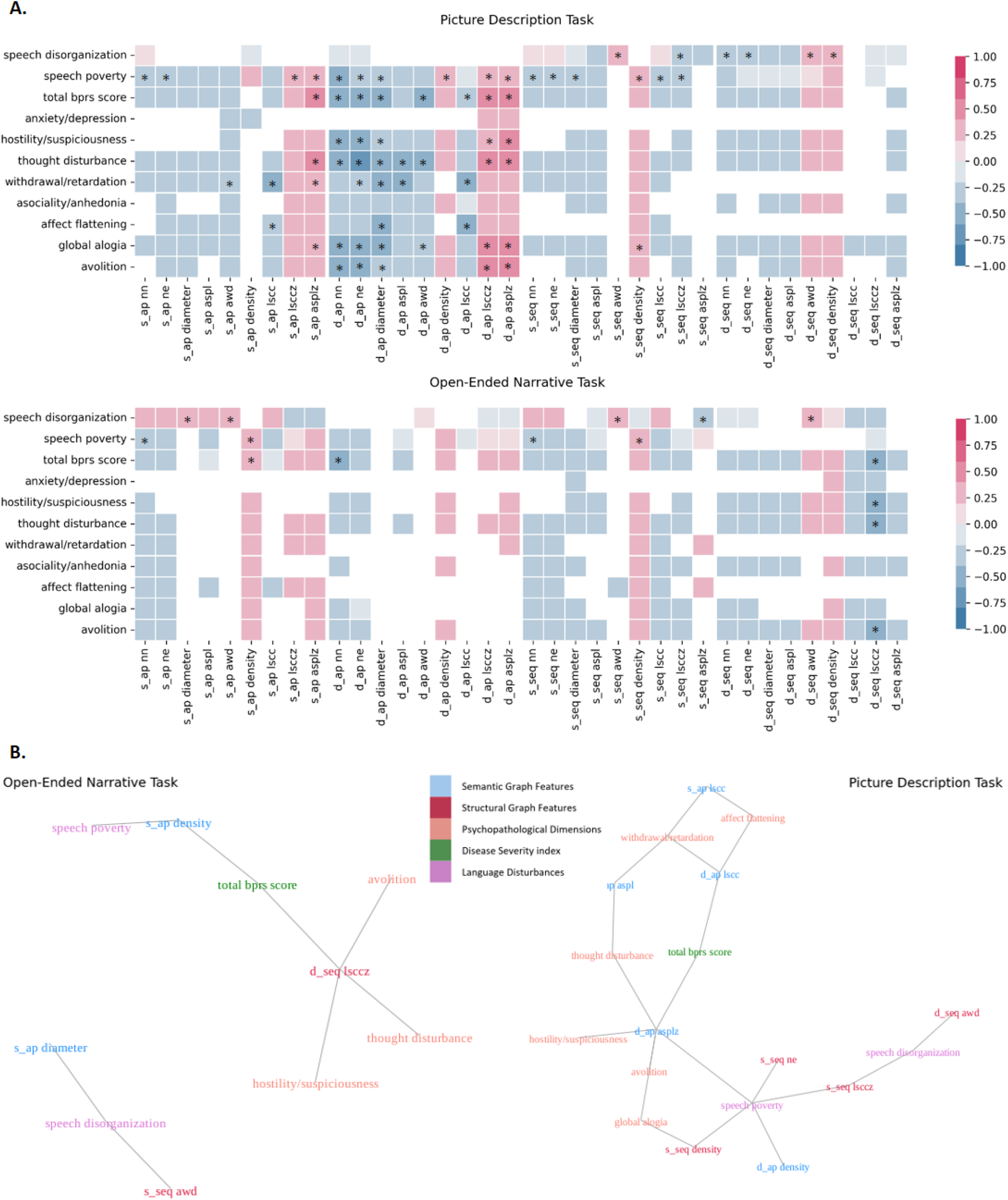
Correlations between structural and semantic graph features and dimensional clinical characteristics. (A) Heatmap representations of the Spearman’s correlation coefficient for structural and semantic graph features and clinical measures in picture description and open-ended narrative tasks across all participants. Significant relationships with uncorrected p values <0.05 are shaded based on their effect sizes (Spearman’s r). Correlations surviving Bonferroni correction are starred. (B) Network representation of significant relationships between graph features and clinical measures. Multi-collinearities were separately handled for structural and semantic graph features by stepwise comparison of variance inflation factors and feature exclusion. Multiple comparisons were accounted for using Bonferroni correction. Note: S_AP = static action-predication graph feature; D_AP = dynamic action-predication graph feature; S_SEQ = static sequential graph feature; D_SEQ = dynamic sequential graph feature; NN = number of nodes; NE = number of edges; Diameter = graph diameter; ASPL = average shortest path length; AWD = average weighted degree; Density = graph density; LSCC = size of largest strongly connected component; LSCCZ = z-score of LSCC compared to 1000 random graphs; ASPLZ = z-score of ASPL compared to 1000 random graphs. More details on graph features are available in Supplemental Table 2.

#### Clinical Measures of Speech Disturbance

Speech disorganization was associated with increased connectedness and decreased organization in both tasks. This relationship is more prominent in structural features with more Bonferroni survived correlations and higher correlation coefficients (absolute r = 0.36 to 0.39). Speech poverty was correlated with density and size of graphs of both type, with impoverished speech having denser and smaller graphs; this correlation was more clearly observed in the semantic networks than in the structural graphs (absolute r = 0.40 to 0.41).

#### Overall Disease Severity

The strongest correlates of total BPRS score were dynamic measures of size (r = −0.54 to −0.58) and organization (r = 0.51 to 0.54) of semantic graphs in picture description task. For narratives, the same relationships were reproduced, but there was a stronger correlation between overall disease severity (BPRS total score) and the dynamic organization measure (LSCCZ) of structural graphs (r = −0.45).

#### Dimensional Measures of Psychosis

Figure 2-B shows the significant relationships among non-redundant graph features and clinical measures. Patterns were task-dependent. Semantic graph features derived from picture description tasks were consistently more informative for dimensional measures of psychosis than structural features. Multiple connections were observed between semantic graph features from picture description and multiple psychopathological dimensions including hostility/suspiciousness, thought disturbance, withdrawal/psychomotor retardation, affective flattening and avolition. Thought disturbance was strongly correlated with almost all dynamic graph features in the Action-Predication network (r = −0.4 to −0.62). Negative factors of withdrawal/psychomotor retardation and affect flattening were connected to static and dynamic measures of connectedness in semantic graphs. Dynamic measure of structural graph organization (LCCZ) was connected with multiple clinical dimensions in open-ended narrative task, including avolition, thought disturbance and hostility/suspiciousness (absolute r = 0.4 to 0.45). The only domain that remained uncorrelated in both task for all graph features was the anxiety/depression factor.

## DISCUSSION

Here, we present a novel and informative automated speech analysis method which objectively measures speech quantity, connectedness and organization using graph metrics. These features are quantitative and objective, and they capture semantic relationships of action (between actors and undergoers of utterance) and predication (between the predicate and arguments).

We found that the semantic graph features derived from the picture description tasks were strongly correlated with psychopathological dimensions of psychosis, with the dynamic features outperforming the static features. Our findings suggest that incorporating semantic information in graph modelling of speech can increase the performance of such models. In the picture description tasks, only the semantic graph features were connected to psychopathological dimensions other than language disturbances, including dimensions such as affect, avolition and thought content. There are no other existing reports of the use of semantic graph methodology in studying psychosis. Therefore, this finding remains to be replicated.

Speech graphs of PS+ were smaller, less connected, and more randomly organized compared to those of PS-. This was true for both semantic and structural graphs in our study, and the finding is consistent with previous studies using structural speech graph [11-13]. For example, Mota et al. found that schizophrenia was related to decreased size in terms of number of nodes [11] and measures of internal distances [12]. The association between psychosis speech and decreased graph size may reflect a similar phenomenon as the decreased semantic density found by Rezaii et al [5]. With regard to measures of connectedness and organization, recent studies have relied on the feature LSCC, the largest strongly connected component, as an absolute and relative quantity with respect to randomly generated graphs [8, 19, 20]. Our findings suggest that additional information can be captured with other non-redundant features describing size, connectedness and organization – for example, we found that diameter and average short path length convey non-redundant information about the expansiveness of the network in addition to number of nodes and edges, network density and average degree can be used along the connectedness measure of LSCC, and z-score analysis of ASPL can be considered as a measure of speech graph organization. Each of these graph metric domains should be included in future efforts to quantify clinical speech characteristics.

We found that graph features are related to psychosis in a task dependent manner. Previous studies reported that dream reports and picture description tasks are more informative about psychosis compared to narrations pertaining to everyday life [12, 13]. Since not all participants are able to recall and report dreams [13], picture descriptions have developed into the preferred source of speech samples for graph analysis [8, 19]. Accordingly, our findings suggest better performance of speech graphs based on picture description tasks compared to open-ended narratives. It may be that the additional structure provided in these tasks is able to reduce noise – i.e., variations in speech that are not related to psychopathology, which may be more dependent on mood or social status of individual speakers. Furthermore, relative to open-ended narratives, picture description tasks have a pre-set common ground between speaker and listener: the picture. This setting helps navigate the speaker’s response, making it *relevant* and *informative* enough for graph analysis.

Moreover, we found that controlling the amount of speech using moving-window method enhanced the performance of speech graph models. This is in line with previous studies conducted by structural speech graphs, where different approaches attempted to control speech quantity, including using normalized features (i.e. measuring features per word count) [11], setting time limits for speech recordings [8, 13, 19], and incorporating moving-window methods (i.e. using dynamic graph features) [8, 12, 13]. Mota and colleagues reported that measuring graph features per word count reversed the relationship of graph features with psychopathological conditions (e.g., the number of nodes were lower in the speech graph of patients with schizophrenia compared to that of patients with manic disorder, but this was reversed when normalized for word count) [11]. Therefore, to make the methodology more uniform and applicable to variety of samples, we suggest using dynamic graph features for subsequent studies.

There are several limitations to the current study. The scope of our comparisons was focused on psychosis as a heterogeneous and multi-faceted condition. Future studies should evaluate whether there are more fine-grained relationships between semantic graph features and psychosis subtypes, as well as whether the presence of potentially comorbid conditions, differences in treatment history, and social determinants affect these measures. Sampling methods were not uniform across the entire sample, as further detailed in the method and supplement. Our primary findings remained consistent when accounting for these deviations statistically. Although we utilized lemmatization of words to merge different inflected forms of lexical units, shared entities that are addressed by different words are stayed detached from each other. Incorporating an algorithm to identify co-referents can help better represent the semantic structure of discourse. We have limited our semantic model to the relations between actors and undergoers. However, semantic theories have identified a variety of more differentiated semantic roles such as experiencer, instrument, source and goal that can be used to produce more fine-grained semantic representations of speech.

The ultimate goal of computational speech measures in the context of psychosis is to develop scalable quantitative methods that improve our understanding of the psychosis disease process and improve our ability to deliver the right treatment to the right person at the right time. The development of novel and informative computational methods moves the field closer to these goals. Our work suggests that graphical speech measures based on semantic relationships capture unique and clinically meaningful aspects of psychosis-related speech disturbances. This novel method was particularly informative when combined with a moving-windows technique and semi-structured tasks. Future efforts may further refine the semantic graph approach by incorporating more differentiated semantic categories for comprehensive characterization of speech in psychosis spectrum disorders and by relating these features to clinical outcomes like relapse risk and treatment response.

## METHODS

### Data Acquisition and Clinical Assessment

Participants (N = 205) were recruited from the Zucker Hillside Hospital inpatient and outpatient services; healthy volunteers were recruited based on prior participation in other studies or through online advertisements. Primary diagnoses of psychotic disorders were established among 81 individuals (36 schizophrenia, 10 schizoaffective disorder, 5 schizophreniform disorder, 18 unspecified psychotic disorder, 12 mood disorder with psychotic features). 87 had primary diagnoses of non-psychotic conditions, and 37 were healthy volunteers. We were interested in the main effect of psychosis, and so chose to compare PS+ (including schizophrenia spectrum and mood disorders with psychotic features) with PS-participants (healthy volunteers and individuals with non-psychotic disorders). All procedures were approved by the Institutional Review Board and all participants provided informed consent or assent as minors. Each participant provided speech samples in response to three picture description tasks and two open-ended narrative prompts. The speech samples were collected via three sampling protocols with different ascertainment goals and minor differences in procedures, as detailed in Supplemental Table 1. All assessments and rating scales were completed by the same team of trained research coordinators (SB, LB). Statistically accounting for the protocol differences did not change our primary findings. Additional details on participants and methods are provided in the Supplement.

Clinical measures included the Scale for Assessment of Thought, Language and communication (TLC) for speech and language disturbances [21], the Scale for Assessment of Negative Symptoms (SANS) [22], and the Brief Psychiatric Rating Scale (BPRS) for overall psychosis symptoms [23]. Two factor scores were calculated from the TLC based on the factor model by Peralta et al: speech disorganization (pressure of speech, tangentiality, derailment, incoherence, illogicality, circumstantiality and loss of goal) and speech poverty (poverty of speech and poverty of content) [24, 25]. Global scores were taken from the SANS for affective flattening, alogia, avolition, and asociality/anhedonia domains of negative symptoms. BPRS total scores were used as a measure for general psychopathology, in addition to its four factors describing anxiety/depression, hostility/suspiciousness, thought disturbance and withdrawal/psychomotor retardation [23].

### Language Pre-Processing

Utterance boundaries were determined manually based on syntactic completeness and the presence of pauses. Within each utterance, tokens were identified by NLTK word-tokenizer [26]. Each token was tagged for its part-of-speech (POS) and lemmatized using spaCy modules [27]. Semantic role labelling (SRL) was performed using transformer-srl (https://github.com/Riccorl/transformer-srl), a BERT-based model built as an extension to AllenNLP and pre-trained on CoNLL 2012 dataset derived from the OntoNotes v5.0 corpus [28-32].

For structural graphs, we included nouns, pronouns, non-auxiliary verbs, adjectives and adverbs. Interjections, filled pauses, articles and conjunctions (e.g., “yes,” “um,” “and then”) were excluded so as to capture tokens that contribute to verbal exchanges. The lemmatized forms of the included tokens were passed to the graph formation step. We chose to lemmatize the tokens to avoid dissociation of different morphological forms of the same lexeme in network representations (e.g., *talk, talked* and *talking* all merged into the same node labeled as *talk*). For action-predication graphs we tagged verbs, actor-arguments and undergoer-arguments using SRL, and the lemmatized forms of them were passed to the graph formation step.

### Statistical Analysis

The associations between graph features and psychosis diagnosis as a dichotomous variable were evaluated using the Mann-Whitney U test. Rank biserial correlation coefficients (RBC) were used as measures of effect size [33]. Graph features were correlated with dimensional clinical measures using Spearman’s rank-order test. The significance threshold was adjusted by Bonferroni correction (adjusted α= 0.0001). The correlations between graph features corresponding to each task and clinical measures were plotted as a heatmap.

Variance inflation factor (VIF) comparison was used to identify redundant graph features in order to simplify our analysis and generate a more streamlined set of features for future applications. First, we applied the method to different features within each domain (size, connectedness and organization) for static and dynamic types separately. Features with the highest VIF were removed successively until all features showed the VIF of less than 5 [18]. The remaining static and dynamic graph features were integrated for each graph type and the same analyses were conducted. These remaining graph features for structural and semantic graphs were then combined to produce non-redundant graph feature sets for each graph type. A final set of graph features were attained by merging all previously survived features and re-doing VIF analysis over them. The significant relationships between the ultimately survived features and clinical measures were demonstrated as a network for each task.

The potentially confounding effect of different sampling protocols on our results was evaluated by covarying for protocol type in multiple linear regressions predicting clinical measures with graph features of interest (survived VIF and Bonferroni tests). All initial findings remained significant.

All statistical analyses and visualizations were done in python using the Pandas [34], NumPy [35], Pingouin [36], SciPy [37], Statsmodels [38], Seaborn [39], igraph [16], and Matplotlib [40] libraries.

## Data Availability

The extracted features and clinical ratings for the participants are provided at https://github.com/STANG-lab/Semantic-Graphs. De-identified raw transcripts can be made available to interested scientists upon request.

## CODE AVAILABILITY

The codes used for language processing, graph formation, and the statistical analyses are available in a public repository at https://github.com/STANG-lab/Semantic-Graphs.

## ACKNOWLEDGEMENTS

This project was supported by the Brain and Behavior Research Foundation Young Investigator Award (SXT) and the American Society of Clinical Psychopharmacology Early Career Research Award (SXT). Data for a portion of the participants (n=132) was collected in partnership with, and with financial support from, Winterlight Labs, Inc. We thank the participants for their contributions. We are also grateful to the clinicians and administrators at Zucker Hillside Hospital for their support of our work, including Drs. Michael Birnbaum, Anna Costakis, and Ema Saito. We thank Bill Simpson, Jessica Robin, and Liam Kaufman of Winterlight Labs for their contributions to the ongoing collaborations.

## AUTHOR CONTRIBUTIONS

Dr. Nikzad designed the study, carried out the data processing and analyses, and drafted the manuscript. Drs. Tang, Cong, Hänsel, Cho, Pradhan, DeSousa and Liberman contributed to the study design and contributed to the manuscript. Ms. Berretta and Ms. Behbehani collected the data and contributed to the manuscript.

## Footnotes

1 Here, we define “speech” as the sum of all acoustic and lexical aspects of spoken communication.

