## Supplemental Tables for "Who Does What to Whom? Graph Representations of Action-Predication in Speech Relate to Psychopathological Dimensions of Psychosis"

|  | Protocol 1  n = 12 | Protocol 1 – Virtual  n = 61 | Protocol 2  n = 105 | Protocol 3  n = 27 |
| --- | --- | --- | --- | --- |
| Age: mean years (SD) | 25.71 (5.17) | 27.68 (4.99) | 24.06 (6.45) | 25.72 (5.33) |
| Sex: n (%) | Female: 5 (42%)  Male: 7 (58%) | Female: 32 (52%)  Male: 29 (48%) | Female: 72 (69%)  Male: 33 (31%) | Female: 9 (33%)  Male: 18 (67%) |
| Gender: n (%) | Woman: 4 (33%)  Man: 6 (50%)  Genderqueer/Gender non-conforming/non-binary/other: 2 (17%) | Woman: 31 (51%)  Man: 28 (46%)  Genderqueer/Gender non-conforming/non-binary/other: 2 (3%) | Woman: 58 (55%)  Man: 36 (34%)  Genderqueer/Gender non-conforming/non-binary/other: 11 (10%)  Prefer not to answer: 1 (1%) | Woman: 5 (19%)  Man: 18 (69%)  Genderqueer/Gender non-conforming/non-binary/other: 2 (8%)  Prefer not to answer: 1 (4%) |
| Psychotic Spectrum Disorder: n (%)  Break down and comorbidities | Total: 12 (100%)  Schizophrenia: 7 (58%)  Schizoaffective Disorder: 2 (17%)  Unspecified Psychotic Disorder: 3 (25%)  Mood disorder: 5 (42%)  Anxiety disorder: 3 (25%)  Substance use disorder: 6 (50%) | Total: 17 (28%)  Schizophrenia: 9 (53%)  Schizoaffective: 1 (6%)  Schizophreniform Disorder: 2 (12%)  Unspecified Psychosis: 4 (23.5%)  Mood disorder: 6 (35.3%)  Anxiety disorder: 2 (12%)  Substance use disorder: 6 (35%) | Total: 25 (24%)  Schizophrenia: 4 (16%)  Schizoaffective: 3 (12%)  Unspecified Psychosis: 7 (28%)  Mood disorder + psychotic features: 12 (48%) | Total: 27 (100%)  Schizophrenia: 16 (59%)  Schizoaffective: 4 (15%)  Schizophreniform: 3 (11%)  Unspecified Psychosis: 4 (15%)  Mood disorder: 5 (18.5%)  Substance use disorder: 5 (18.5%) |
| Other Psychiatric Conditions: n (%)  Break down and comorbidities | Total: 0 (0%) | Total: 44 (72%)  Healthy Volunteer: 37 (84%)  Anxiety disorder: 7 (16%)  Mood disorder: 2 (4.5%)  Substance use disorder: 3 (7%) | Total: 80 (76%)  Mood disorder: 76 (95%)  Anxiety disorder: 19 (24%)  Substance use disorder: 12 (15%) | Total: 0 (0%) |
| Ascertainment | Inpatient | Outpatient and healthy volunteers | Inpatient and outpatient | Inpatient |
| Data Collection Method | Recorded in-person interaction | Recorded interaction over Microsoft Teams | In-person data collection proctored with digital app | In-person data collection proctored with digital app |
| Assessment Context | Other assessments included semi-structured clinical interviews and self-report scales.  Scales included in this analysis: BPRS, SANS, TLC. | Other assessments included semi-structured clinical interviews and self-report scales.  Scales included in this analysis: BPRS, SANS, TLC. | Other assessments consist of self-report questionnaires.  Scales included in this analysis: TLC. | Other assessments included semi-structured clinical interviews  Scales included in this analysis: BPRS, SANS, TLC. |
| Picture Description Stimuli | 1. Cookie theft scene  2. Rorschach  3. TAT | 1. Cookie theft scene  2. Rorschach  3. TAT | 1. Family kitchen scene  2. Rorschach  3. TAT | 1. Lightbulb living Room scene  2. Rorschach  3. Family kitchen scene |
| Open-ended Narrative Prompts | 1. Tell me about yourself  2. How have things been going recently? | 1. Tell me about yourself  2. How have things been going recently? | 1. Tell me about yourself  2. How have you been spending your time, recently? | 1. Tell me about yourself  2. How have you spent your time recently? |

**Supplemental Table 1.** Comprehensive clinical characteristics of participants. Note: BPRS = Brief Psychiatric Rating Scale; SANS = Scale for Assessment of Negative Symptoms; TLC = Scale for the Assessment of Thought, Language and Communication; TAT = Thematic Apperception Test; SD = Standard Deviation.

| Graph Feature | Symbol | Definition | Domain |
| --- | --- | --- | --- |
| Number of Nodes | NN | Number of distinct nodes | Size |
| Number of Edges | NE | Number of distinct edges | Size |
| Diameter | Diameter | Shortest path length between the two most distant nodes | Size |
| Average Shortest Path Length | ASPL | Average number edges to pass from one randomly selected node to another | Size |
| Average Weighted Degree | AWD | Average sum of weights of edges per node | Connectedness |
| Density | Density | Number of realized edges divided by number of possible edges | Connectedness |
| Size of Largest Strongly Connected Component | LSCC | The size of largest component of the graph with all nodes being mutually reachable | Connectedness |
| The z-score of the Size of Largest Strongly Connected Component compared to 1000 random graphs | LSCCZ | z-score of LSCC in the population of 1000 randomly generated graphs of the same size (similar NN and NE) | Organization |
| The z-score of the Average Shortest Path Length compared to 1000 random graphs | ASPLZ | z-score of ASPL in the population of 1000 randomly generated graphs of the same size (similar NN and NE) | Organization |

**Supplemental Table 2.** Symbols and definitions of graph features.
